# Performance of an Ambient Generative AI Documentation Tool in a Linguistically Diverse Clinical Setting

**DOI:** 10.64898/2026.08.14.26360467

**Authors:** Rajendra Aldis, Serena Wang, Michael Sage, Michael Metzmaker, Hannah Galvin

## Abstract

Ambient artificial intelligence scribes are being increasingly used in healthcare to improve efficiency and reduce provider clinical documentation burden, yet their performance across linguistically diverse patient populations is not well characterized. We conducted a retrospective analysis of 54,160 outpatient encounters within a U.S. safety net health system to evaluate the performance of an artificial intelligence documentation tool in English and non-English clinical encounters, and in encounters where an interpreter or bilingual provider was present. Documentation performance was measured by the percentage of words in the final note that were generated by the ambient AI documentation tool and not edited by the provider. Associations between language factors and documentation performance were measured using Generalized Estimating Equations with exchangeable correlation structures to account for clustering of multiple encounters within unique patients. Univariable models were fitted to estimate the odds of adequate performance by language and interpreter modality, and a multivariable interaction model was used to evaluate within-language differences between bilingual providers and interpreter-mediated encounters. Non-English encounters were 21%–25% less likely than English encounters to achieve the same performance threshold. There was no significant difference in generative documentation performance between interpreter-mediated and bilingual provider encounters. These findings underscore the importance of equity-focused evaluation and multilingual model refinement to ensure that artificial intelligence documentation benefits are distributed fairly across diverse patient populations.

## Introduction

The introduction of ambient artificial intelligence (AI) scribes into healthcare environments represents a fundamental shift in how providers document clinical care. By combining automatic speech recognition (ASR), speaker diarization, natural language processing (NLP), and large language models (LLMs), ambient scribes passively capture conversational dialogue during clinical encounters and automatically generate structured clinical notes. Early pragmatic evaluations have demonstrated significant benefits, including a 13-percentage-point reduction in burnout prevalence and a 20% to 30% decrease in documentation time.^1-4^ Ambient AI scribes have also been shown to restore the focus of the clinical encounter to the patient, as providers no longer need to prioritize the keyboard over face-to-face interaction.^5^

However, the performance of these tools in safety net settings and among linguistically diverse populations remains understudied. Existing studies either relied on synthetic or controlled dialogues rather than real world clinical care, or failed to examine variation by patient language or interpreter involvement.^6-11^ Safety net systems serve populations that face significant systemic barriers, including limited English proficiency, which necessitates use of professional interpreters or care by bilingual providers. There is concern that AI tools can exhibit lower accuracy or utility in these settings,^12-14^ risk creating a documentation tax for providers serving marginalized populations, and potentially create or exacerbate health disparities.^15-17^ Additionally, inaccurate AI documentation can have downstream knock on effects; because clinical notes serve as the primary communication vector across multidisciplinary teams, uncorrected omissions or factual mistakes in ambient documentation become part of the permanent longitudinal EHR once signed, endangering patient safety by potentially introducing erroneous information into subsequent clinical decision-making.^18^ If ambient AI documentation performance is distributed unequally across language groups, vulnerable populations may be left exposed to greater downstream risk.

These potential disparities are rooted in established architectural limitations of current ambient AI documentation systems. Automatic speech recognition algorithms, the foundational layer of ambient listening, exhibit significantly higher word error rates for speakers with non-native accents or distinct dialects due to their underrepresentation in training datasets.^19,20^ Additionally, standard ambient models are optimized for dyadic (physician-patient) interactions, while the introduction of an interpreter creates a complex triadic workflow complicated by translation lag and overlapping speech, which challenges the speaker diarization algorithms required to accurately attribute text to the correct participant.^21,22^ If the model fails to parse these interactions, the burden of synthesizing the clinical note reverts to the provider, negating the tool’s intended efficiency gains.

This study utilizes data from a safety net healthcare system to evaluate the performance of an ambient AI documentation tool for a linguistically diverse population. By analyzing over 54,000 clinical encounters, we investigate whether documentation performance disparities exist between English and non-English clinical encounters. We also assess whether the mode of language assistance, specifically the use of interpreters versus certified bilingual providers, affects ambient AI documentation performance. This research seeks to contribute to the emerging body of knowledge on the implementation of generative AI in healthcare, emphasizing the necessity of equity and linguistic inclusivity in the development and implementation of generative AI based technologies.

## Methods

### Study Design and Setting

We conducted a retrospective cross-sectional study of outpatient clinical encounters that occurred in an academic safety net health system that serves approximately 140,000 unique patients annually across the greater Boston metropolitan area. The patient population is characterized by a high degree of linguistic diversity; over half the patients speak a language other than English at home, and 42% of primary care patients have limited English proficiency and need a professional medical interpreter. The system provides face-to-face, telephonic and video conference interpreter services in over 60 languages. Clinical encounters are conducted both in-person and virtually. The project was reviewed by the CHA Institutional Review Board and received a determination of Not Human Subjects Research.

### Intervention and Workflow

The health system began to implement an ambient generative AI documentation tool (Abridge) in March 2024. The tool utilizes a multi-stage machine learning pipeline: The initial stage uses ASR to convert acoustic signals into a written transcript, which includes speaker diarization to distinguish between the clinician, the patient, and any third parties (such as family members or professional interpreters). A generative LLM then extracts clinically relevant entities from the transcript and synthesizes them into structured clinical note sections, such as history of present illness, and assessment\plan. The pipeline can generate an English clinical note from encounters conducted in 28 different languages.

The tool was implemented in a phased roll out, with the initial cohort including physicians and advanced practice providers in primary care, pulmonary medicine, and orthopedics.

Subsequent phases occurred over 18 months and included additional specialities such as surgery, ophthalmology, podiatry, dermatology, cardiology, neurology, and obstetrics/gynecology. Participating providers completed standardized training on ambient dictation functionality and workflow integration.

During encounters, providers enabled the ambient listening documentation tool on their smartphone using either the vendor’s application or the EHR mobile application. Encounters could be in-person or televisits. Verbal consent was obtained from patients prior to recording. For encounters involving non-English languages, providers utilized standard interpreter workflows (either professional remote interpreter or in-person interpreter) or conducted the visit as a certified bilingual provider; no specific modifications to the AI workflow were required for these scenarios.

To initiate ambient listening, providers tapped an icon in the application, and when the visit was completed they tapped another icon to generate the note. After initiation, the tool recorded ambient audio of the conversation between the provider, patient, and, if present, the interpreter. After recording termination, the tool generated a draft note typically within 30 seconds and made the note available for review and editing directly within the EHR. Providers were also able to view the transcript of the encounter that the tool used to create the note draft. Ground truth audio records were also available. Providers retained full discretion to review, edit, accept, or reject the AI-generated text to ensure accuracy.

### Data Sources and Variables

Primary outcome data was supplied by the vendor via weekly flat file transfers. All other data elements were extracted from the EHR analytic data base. Data for all completed outpatient encounters utilizing the ambient AI tool between March 2024 and August 2025 were included in the analysis.

The primary outcome was Percent Effort Reduction (PER), a vendor-reported metric representing the percentage of words in the final note that were generated by the ambient AI documentation tool and incorporated by the provider as-is. Specifically, PER is defined as

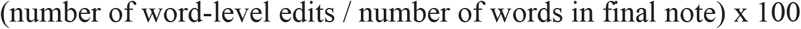

A value of 100 means the providers did not make any edits to the note, and 0 means that the provider rejected the entire note. The metric was generated for the history of present illness and assessment/plan sections of the note, with the average weighted by the number of words in each section.

To evaluate clinical utility, we dichotomized PER at a threshold of *⪰*80%, which aligns with vendor operational benchmarks used to identify sufficient quality generated documents. While prior evaluations of ambient AI have quantified overall adoption using total character counts or subjective quality scores, PER more directly captures word-level clinician modification.^11,23^

Independent variables were 1) patient primary language (English, Spanish, Portuguese, Haitian Creole, Other) and 2) language assistance modality. Language assistance was categorized into three mutually exclusive groups: no interpreter, interpreter used, and certified bilingual provider.

### Statistical Analysis

We modeled the association between language factors and PER using Generalized Estimating Equations (GEE) with a binomial distribution and logit link function. An exchangeable correlation structure was used to account for clustering of multiple encounters within unique patients. Univariable models were first fitted to estimate the odds of optimal performance by language and interpreter modality. To evaluate within-language differences between bilingual providers and interpreter-mediated encounters, a single pooled GEE model was constructed across all non-English encounters. Within-language contrasts comparing bilingual provider visits to interpreter-mediated visits (e.g., Spanish bilingual vs. Spanish interpreter) were evaluated using linear contrast Wald tests on the model parameters. A full-interaction model was also fitted and omnibus Type III Wald tests were used to measure the overall interaction effect. Results are reported as odds ratios (OR) with 95% confidence intervals (CI). All statistical tests were two-tailed with alpha set to 0.05. *P* values are reported to three decimal places. Analyses were performed using Python version 3.8, utilizing the pandas library for data management and statsmodels (v0.13.2) for GEE implementation.

## Results

### Study Cohort Characteristics

The analytic cohort was composed of 54,160 outpatient clinical encounters conducted between March 2024 and August 2025. The majority of encounters were conducted in English (n=39,478; 72.9%). Among encounters conducted in non-English languages, Portuguese was the most prevalent language (n=6,380; 11.8%), followed by Spanish (n=4,541; 8.4%), and Haitian Creole (n=2,349; 4.3%). Across all languages, 23.1% (n=12,522) encounters utilized an interpreter, while 4.0% (n=2,161) were conducted by a certified bilingual provider without an interpreter. The distribution of clinical specialties was diverse, with the highest volumes in Family Medicine (29.5%) and Internal Medicine (15.8%) (Table 1.)

**Table 1:** Encounter Characteristics.

|  | n | % |
| --- | --- | --- |
| **Patient Language** |  |  |
| English | 39478 | 72.9 |
| Portuguese | 6380 | 11.8 |
| Spanish | 4541 | 8.4 |
| Haitian Creole | 2349 | 4.3 |
| Other | 1413 | 2.6 |
| **Language Assistance** |  |  |
| No interpreter | 39478 | 72.9 |
| Interpreter | 12522 | 23.1 |
| Bilingual Provider | 2161 | 4.0 |
| **Clinical Specialty** |  |  |
| Family Medicine | 15974 | 29.5 |
| Internal Medicine | 8564 | 15.8 |
| Podiatry | 5941 | 11.0 |
| Pediatrics | 4006 | 7.4 |
| Dermatology | 3606 | 6.7 |
| Orthopedic Surgery | 2522 | 4.7 |
| Other | 13548 | 25.0 |
| **Language Assistance By Language** |  |  |
| Portuguese_Interpreter | 6255 | 11.5 |
| Portuguese_Bilingual | 125 | 0.2 |
| Spanish_Interpreter | 3355 | 6.2 |
| Spanish_Bilingual | 1186 | 2.2 |
| Haitian Creole_Interpreter | 1525 | 2.8 |
| Haitian Creole_Bilingual | 824 | 1.5 |
| Other_Interpreter | 1387 | 2.6 |
| Other_Bilingual | 26 | 0.0 |

### Unadjusted Ambient Documentation Performance

Unadjusted analyses (Table 2), defined as achieving a PER of *⪰*80, revealed that English-language encounters achieved the threshold in 57.6% of cases. In contrast, success rates were lower for all non-English languages: Spanish (51.1%), Portuguese (51.0%), Haitian Creole (50.5%), and Other languages (48.7%). Encounters requiring language assistance showed lower unadjusted performance rates compared to unassisted encounters (57.6%), with minimal difference observed between visits utilizing an interpreter (50.7%) and those conducted by a bilingual provider (50.5%).

**Table 2:**
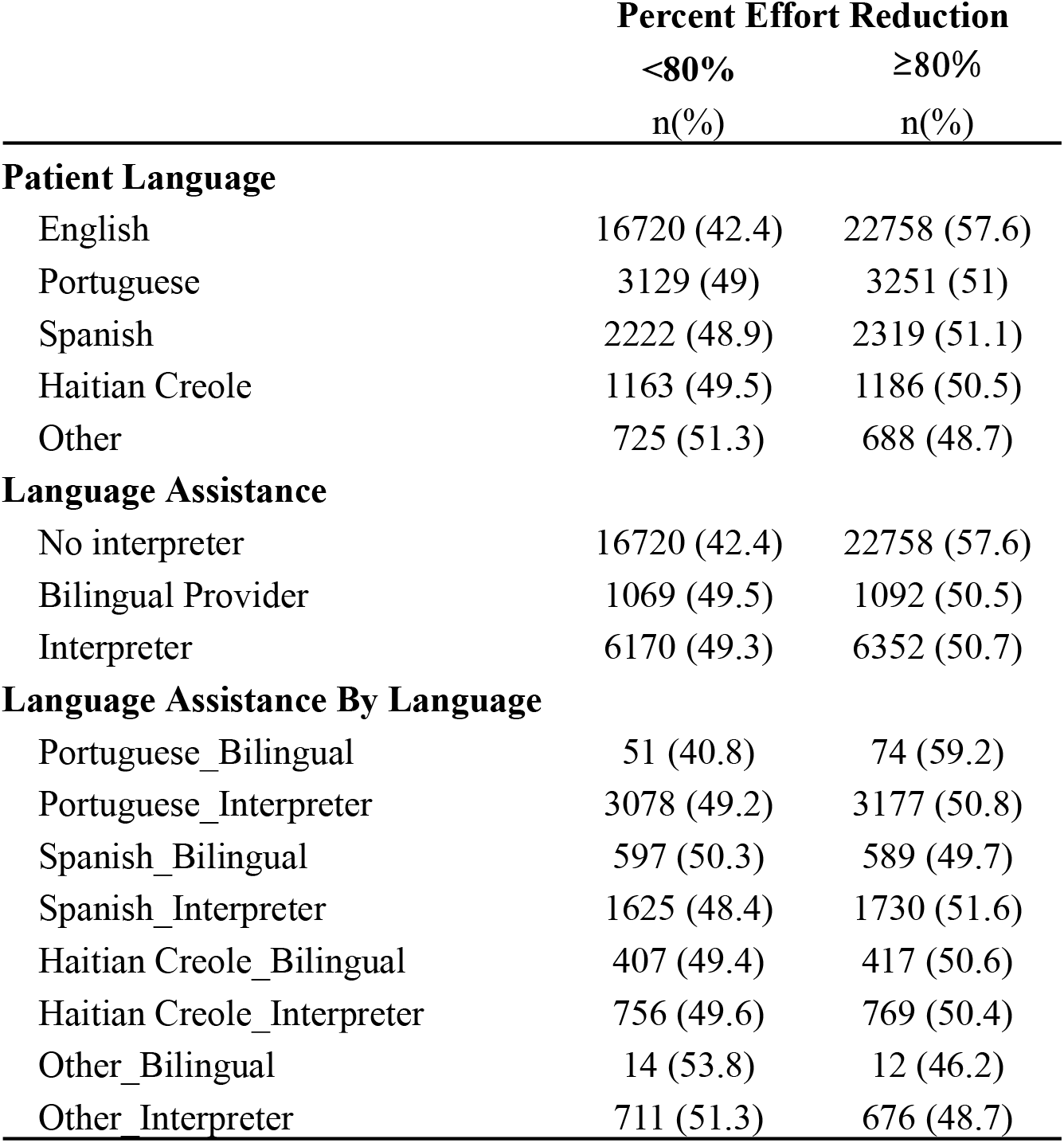
Unadjusted Percentage Effort Reduction Comparison.

### GEE Analysis of Language and Language Assistance Modality

In GEE models patient language was a statistically significant predictor of ambient documentation performance (Table 3). Compared with English-speaking encounters, all non-English language groups had significantly lower odds of achieving *⪰*80 PER. Specifically, encounters conducted in Spanish (OR, 0.76 [95% CI, 0.71–0.81]; *p* = 0.000), Portuguese (OR, 0.79 [95% CI, 0.74–0.83]; *p* = 0.000), and Haitian Creole (OR, 0.75 [95% CI, 0.69–0.82]; *p* = 0.000) were associated with a 21% to 25% reduction in the odds of documentation success. Analysis of language assistance modality demonstrated similar trends. Compared with encounters requiring no interpreter (reference group), both encounters utilizing an interpreter (OR, 0.77 [95% CI, 0.74–0.80]; *p* = 0.000) and those conducted by a bilingual provider (OR, 0.73 [95% CI, 0.67–0.81]; *p* = 0.000) had significantly lower odds of achieving the performance threshold.

**Table 3:** GEE Models of Ambient Dictation ≥80% Effort Reduction.

|  | **OR** | **CI** | ***p*** |
| --- | --- | --- | --- |
| **Patient Language (Ref: English)** |  |  |  |
| Portuguese | 0.79 | [0.74, 0.83] | 0.000 |
| Spanish | 0.76 | [0.71, 0.81] | 0.000 |
| Haitian Creole | 0.75 | [0.69, 0.82] | 0.000 |
| Other | 0.7 | [0.63, 0.79] | 0.000 |
| **Language Assistance (Ref: No Interpreter)** |  |  |  |
| Interpreter Used | 0.77 | [0.74, 0.80] | 0.000 |
| Bilingual Provider | 0.73 | [0.67, 0.81] | 0.000 |
| **Language Assistance by Language (Ref: Interpreter)** |  |  |  |
| Portuguese Bilingual | 1.21 | [0.819, 1.792] | 0.336 |
| Spanish Bilingual | 0.91 | [0.786, 1.060] | 0.230 |
| Haitian Creole Bilingual | 1.00 | [0.834, 1.205] | 0.977 |
| Other Bilingual | 1.00 | [0.488, 2.050] | 0.999 |

### Within-Language Comparison of Language Assistance Modality

To determine whether direct provider-patient communication mitigated performance gaps observed in non-English encounters, we evaluated the impact of bilingual provider status versus interpreter use within specific languages. In this analysis, no statistically significant difference in ambient documentation performance was observed between encounters with bilingual providers and those with interpreters for any language. For Spanish-speaking patients, the odds of success for bilingual providers were not significantly different from those using interpreters (OR, 0.91 [95% CI, 0.79–1.06]; p = 0.230). Similarly, no significant differences based on provider bilingual status were found for Portuguese (OR, 1.21 [95% CI, 0.82–1.79]; p = 0.336), Haitian Creole (OR, 1.00 [95% CI, 0.83–1.21]; p 0.976), or Other languages (OR, 1.00 [95% CI, 0.49–2.05]; p 0.979)

## Discussion

In this large, real-world evaluation of over 54,000 outpatient encounters within a linguistically diverse safety net health system, we identified a consistent decrease in ambient AI documentation performance in encounters conducted in languages other than English: Non-English encounters were 21%–25% less likely than English encounters to achieve ≥80 PER. While moderate in magnitude, this gap was consistent across Spanish, Portuguese, Haitian Creole, and other languages, and demonstrates that even in an urban health system with established interpreter workflows and bilingual provider certification processes, ambient generative AI tools do not yet perform equivalently across all languages. Multilingual LLMs themselves and automatic speech recognition systems have demonstrated reduced performance in underrepresented languages and dialects, frequently attributed to imbalances in training data.^24^ While the comparable performance observed across commonly spoken languages (Spanish, Portuguese) and a language of lesser diffusion (Haitian Creole) suggest baseline multilingual capability, parity with English is lacking.

From an equity perspective, these findings are consequential. Prior studies have shown that ambient AI documentation can save providers millions of keystrokes over time, and lead to decreased provider time spent completing notes.^11,25,26^ Safety net providers disproportionately care for patients with limited English proficiency. If ambient AI tools yield smaller efficiency gains in non-English encounters, providers serving these populations may realize fewer of these workflow benefits. Along with losing efficiency gains, providers seeing patients who speak languages other than English can face the additional cognitive workload of having to find portions of the generated note that need improvement and edit them. Over time, unequal distribution of AI-enabled efficiency gains could reinforce existing disparities in provider workload and burnout risk.

There is an additional concern that the integration of ambient generative AI scribes into routine clinical workflows can introduce automation bias, which is the tendency for clinicians to rely on automated cues as a heuristic replacement for active verification. This form of bias is a well-recognized hazard of interacting with AI-driven health technologies.^27^ Human factors research demonstrates that automation bias is strongly exacerbated under conditions of elevated cognitive load, occurring most frequently when clinicians operate under time constraints or heavy administrative demands that impair their capacity to scrutinize system outputs.^28^ In ambient documentation workflows, automation bias resulting from increased cognitive load due to decreased ambient documentation performance can lead to uncritical acceptance of generated draft notes and allow for latent errors such as factual omissions, hallucinated findings, or diagnostic misattributions to enter the permanent medical record. Once signed, these errors can propagate across multidisciplinary care teams, leading to downstream clinical consequences, including delayed diagnoses, inappropriate pharmacotherapy, and compromised treatment planning.^18^ Because ambient tools demonstrate lower documentation performance in non-English encounters, non-English speaking patients may be at greater risk for these types downstream errors.

The comparable relative performance of interpreter-mediated encounters and those conducted by bilingual providers suggests that the tool successfully managed the additional complexity of triadic communication present in interpreter-mediated encounters, which forces the model to contend with overlapping speech and speaker attribution. This finding also indicates that the presence of an intermediary layer of English provided by an interpreter does not overcome the system’s underlying difficulty in processing non-English clinical dialogue. Consequently, achieving equitable performance in ambient AI documentation will require improvements to how these models process diverse, multilingual conversational audio, rather than relying on interpreter support to compensate for algorithmic limitations.

Despite the current performance disparities, the comparable outcomes observed between bilingual and interpreter-mediated encounters point toward a potential for ambient AI to provide interpretation as well as documentation. The baseline ability of the tool to generate structured clinical notes from encounters conducted entirely in non-English languages suggests that these systems possess an inherent multilingual capability that, with further development, could allow for the integration of real-time, healthcare-specific translation directly into the ambient documentation workflow. Achieving this would require additional fine-tuning on diverse training data sets, and ongoing assessment for algorithmic bias.

Several limitations affect the generalizability and interpretation of our findings. This was a single-center study using a vendor-specific solution, so results may not generalize to other institutions or technologies. Although providers used the ambient tool on both iOS and Android platforms, only the iOS version of the tool was fully integrated with our EHR. As a result, language assistant data could only be linked for EHR integrated encounters, so those conducted via Android were excluded from analysis, potentially introducing platform-related bias. Use of the tool was voluntary for both providers and patients, which could introduce selection bias as there may have been differences in patients willing to consent to AI recording and providers inclined to adopt new technology compared to those who did not. Our primary outcome, PER, is a vendor-defined metric that measures textual preservation within selected note sections. While operationally clear, PER does not directly measure clinical accuracy, completeness, or documentation quality. Finally, we did not control for unmeasured confounders such as visit complexity, provider editing behavior, acoustic environment, provider specialty, or patient case mix. Although the overall sample size was substantial, subgroup analyses for less common languages and specific interpretation modalities may have been underpowered.

Our findings lay the groundwork for future work, which could include incorporating direct measures of documentation accuracy and quality, such as structured manual review or use of validated instruments such as the Physician Documentation Quality Instrument (PDQI-9).^29^ Downstream outcomes, including documentation time, provider burnout, revenue capture, and patient safety, could be evaluated across language strata to determine whether observed differences in effort reduction translate into meaningful experiential or clinical disparities.

Focusing on the different clinical settings, their acoustic properties, and effect on transcription accuracy could inform further ambient dictation development. Conducting these studies across multiple centers would inform generalizability across technologies and practice settings.

Although moderate in magnitude, the performance disparities we found highlight the importance of evaluating generative AI systems in linguistically diverse clinical environments prior to broad deployment. As ambient tools become embedded in routine documentation workflows, systematic assessment across language groups will be necessary to ensure equitable performance. There is also a need for policies that encourage algorithmic transparency, with an emphasis on measuring and reporting algorithmic bias, so health care systems can have a clear understanding of the risks and benefits of implementing an AI-based tool.^30-32^

## Data availability

The datasets generated and/or analyzed during the current study are available from the corresponding author on reasonable request.

## Code availability

The underlying code for this study is not publicly available but may be made available to qualified researchers on reasonable request from the corresponding author.

## Acknowledgements

This project received no-cost biostatistics consultation from the Harvard Catalyst Center. The center receives support from the NIH UM1TR004408 award and financial contributions from Harvard University and its affiliated academic healthcare centers. The content is solely the responsibility of the authors and does not necessarily represent the official views of Harvard Catalyst, Harvard University and its affiliated academic healthcare centers, or the National Institutes of Health.

## Author Contributions

RA participated in conceptualization, methodology, analysis, data curation, writing original draft, review & editing, visualization; SW participated in conceptualization, writing original draft, review & editing; MS participated in conceptualization, methodology, writing; MM participated in conceptualization, writing original draft, review & editing; HG participated in conceptualization, methodology, writing original draft, review & editing, supervision, project administration.

## Human Ethics and Consent to Participate

In accordance with the Declaration of Helsinki, the study was reviewed by the Cambridge Health Alliance Institutional Review Board and received a determination of Not Human Subjects Research. Data used in the study was collected during the course of routine care, and did not require participant consent.

